# Forecasting combination of hierarchical time series: A novel method with an application to COVID-19

**DOI:** 10.1101/2020.10.11.20210799

**Authors:** Livio Fenga

## Abstract

Multiple, hierarchically organized time series are routinely submitted to the forecaster upon request to provide estimates of their future values, regardless the level occupied in the hierarchy. In this paper, a novel method for the prediction of hierarchically structured time series will be presented. The idea is to enhance the quality of the predictions obtained using a technique of the type forecast reconciliation, by applying this procedure to a set of optimally combined predictions, generated by different statistical models. The goodness of the proposed method will be evaluated using the official time series related to the number of people tested positive to the SARS-CoV-2 in each of the Italian regions, between February 24^*th*^ 2020 and August 31^*th*^ 2020.

## 1. Introduction

In many applications, it is often the case that accurate forecasts are needed for time series showing an inherent hierarchical structure. For example, in economics the forecaster is routinely asked to provide separate forecasts for the industrial production index at the most aggregated level as well as for specific (sub–) classes of economic activities. The estimation of the future demand of domestic tourism usually follows a geographical proximity criterion, based on which the related time series are organized (and predicted) according to homogeneous groups. Sometimes, emergency situations require close monitoring of the spread of a disease not only at a national but also at a regional level, e.g. in order to set up more appropriate countermeasures for elderly and chronically ill people. These are all cases where a single line of hierarchy generates the overall structure of the data which therefore is referred to as “hierarchical time series”. The present paper is concerned with the forecast of such data structures. While on one hand it is always possible to disregard the underlying hierarchical arrangement and thus carry out the prediction exercise considering each time series singularly, on the other hand, by doing so, it is very unlikely for the resulting higher level forecasts to be equal to the sum of the lower level ones. It goes by itself that such a situation is not acceptable in many instances, e.g. in the field of official statistics where aggregation consistency is generally a *conditio sine qua non* and even slight misalignment need to be dealt with. Therefore, many combination techniques have been designed to preserve the needed adding-up conditions, by accounting for the position occupied in the hierarchy by each and every time series, regardless their level of (dis–)aggregation. However, this type of approach – usually referred to as forecast reconciliation – is in general dependent on the statistical model *a priori* chosen to carry out the forecasting exercise. Undoubtedly, this choice might negatively impact the quality of the generated forecasts, e.g. by conveying not negligible amount of uncertainty into the analysis. This is especially true in the case of real-life data – where problems, such as small sample size, noise and systematic and/or non systematic errors – might hinder the choice of the “right” statistical model.

Motivated by this, in the present paper a method built upon the forecast reconciliation procedure devised by Hyndman, R. J., Ahmed, R. A., Athanasopoulos, G., and Shang, H. L. (2011) will be presented. In more details, it will be formulated a joint hierarchical forecasting system, where an additional optimality condition, derived in a multi-model setup of the type forecast combination, drives the choice of the “best” statistical model generating the predicted values. A final step, designed to lower the bias of the selected forecasts, is also part of the procedure. The main novelty of the method is that the forecast combination is applied directly on forecasts which have already been reconciled. In essence, it is an optimization procedure articulated in four steps: one performed at a cross-section level (reconciliation), two at a cross-model level (forecast combination) and the final one on the chosen prediction vector, for bias adjustment purposes. Surprisingly, to the best of the author’s knowledge, this is the first attempt of this sort in the case of cross-sectional hierarchical time series.

The rest of the paper is structured as follows: Section 2 is devoted to the literature review concerning the two statistical methods the proposed procedure is based on, which will be detailed in the following Section 3. The proposed method, as well as its justification, will be respectively illustrated in Section 4 and 5. The following Section 6 will be devoted to an extensive empirical application, carried out using the official Italian data related to the SARS-CoV-2 positive cases, which will demonstrate the validity of the proposed approach. Section **??**, containing the conclusions and the future directions of this work, will end the paper.

## 2. Literature review

As already mentioned, the proposed procedure is based on two class of methods, usually referred to as forecast reconciliation and forecast combination. The former serves the purpose of achieving aggregation consistency of individual, aggregation inconsistent, forecasts whereas the latter will be employed to combine different reconciled forecasts, each of them generated according to different statistical models.

A rigorous and theoretically sound investigation on forecasts combination dates back to the late 60s – with the famous seminal paper by Bates, J. M., and Granger, C. W. (1969). Here, the Authors showed that the combination of forecasts often leads to a better forecast accuracy and, by doing so, provided an alternative way to the notion that a “best” method exists and can be identified. Ever since this paper, the integration of a number of forecasts, independently estimated on a single time series, has attracted a great deal of research interest and, as a result, a vast literature is today available. Much of it is aimed at presenting empirical applications documenting the appealing features of this approach, which in many cases can improve even upon the best individual forecast, in terms of forecast risk, forecast error variance and consistency between in-sample and out-of-sample error distributions, as pointed out by Barrow, D. K., and Kourentzes, N. (2016). This can happen for a variety of reasons, many of them related to the fact that the choice of the “right” model, in general, implies the injection of not negligible amounts of uncertainty into the analysis (Chambers, J. C., Mullick, S. K., and Smith, D. D. (1971) and Chatfield, C. (1996). In the same line of thinking, many Authors, see for example Makridakis, S. (1989), Stock, J. H., and Watson, M. W (2001) and Stock, J. H., and Watson, M. W. (2004), emphasize the dangers related to misspecification errors, which, on the other hand, can be mitigated by combining the forecasts yielded by a number of models. In support of this argument, there are a number of studies where it is shown that it is very unlikely, using a well calibrated portfolio of models, that one of them consistently dominates the others across the whole prediction window. Such an argument is consistent with the view that the “true” underlying data generating process is, saved for trivial or lab controlled cases, way too complicated to be adequately captured by a single model. This is the position, for example, of Buckland, S. T., Burnham, K. P., and Augustin, N. H. (1997), according to whom the data can never support, and we can never identify, the “true” model. Therefore, the selection of a statistical model is more realistically the process of identifying the best approximating one. Once defined models as approximations, the concept of the identification of the “true” one ceases to be decisive in favor of approaches pursuing, in the first place, the goal of achieving good forecasting performances.

Many practical uses of forecasts combination are discussed in the excellent work of Clemen, R. T. (1989), where the Author covers a wide spectrum of applications, ranging from economics, demography and politics to meteorology and outcomes of football games. In the same spirit is the more recent paper by Mancuso, A. C. B., and Werner, L. (2013), which presents a classification of 174 articles focusing on forecast combination. In particular, new applications are reported from different sectors, such as commercial, tourism, urban traffic, betting market and propagation of successful innovations. The analysis of the outcomes of the M3 forecast competition, discussed in Makridakis, S., and Hibon, M. (2000), goes in favor of the forecast combination approach, which on average proved to outperform the methods individually applied. Same conclusion applies to the more recent M4 forecast competition, discussed in Mancuso, A. C. B., and Werner, L. (2020), where 12 out of a group of 17 most accurate methods are based on the combination of forecasts. Other considerations in favor of this approach are more closely related to the features of the time series under investigation. For instance, in many real-life cases they are affected by structural breaks, induced by a variety of factors whose real-time detection is generally difficult to achieve. However, in a multi–model setup it is not unreasonable to have models showing different degrees of ability in handling such events. Such a situation can translate into gains in terms of forecast accuracy, as it has been argued not only since the very beginning, in the above mentioned paper by Bates and Granger, but also in more recent times, by, among others, Clements, M. P., and Hendry, D. F. (2002), Sessions, D. N., and Chatterjee, S. (1989) and Makridakis, S. (1989). All these Authors concur on the premise that, on average, combining the forecasts yielded by models with different reaction times to a given intervention – and thus requiring stretches of post-break data of different length – can do a better job than individual models. For what said, it comes at no surprise that such appealing results might lead to a change in the perspective many researchers and practitioners look at the forecasting methods, i.e. from model selection – based on the assumption of the existence of one, “true” data generating process – to model averaging.

When the data set under investigation show a hierarchical structure, forecast combination techniques (but this holds true for any univariate forecasting procedure) are insensitive to the level of aggregation at which they are applied. Consistently, in such cases, the independent forecasting of the component time series is always possible, even if not advisable, due to the very likely lack of consistency occurring between the sum of the predictions generated at one level with those available at the level above. But this is not the whole story: by applying forecasting procedures at the components level, rather than limit them to the most aggregate one, it is possible to adequately capture the data covariance structure and thus achieve not negligible gains in terms of quality of the predictions. This fact has been pointed out, *inter alia*, by Fair, R. C., and Shiller, R. J. (1990), Marcellino, M., Stock, J. H., and Watson, M. W. (2003) and Hubrich, K. (2005). Their conclusions are related to two of the most traditional approaches, usually referred to as Top-Down and Bottom-Up (see, for example, Schwarzkopf, A. B., Tersine, R. J., and Morris, J. S. (1988), Lapide, L. (2006) and Athanasopoulos, G., Ahmed, R. A., and Hyndman, R. J. (2009)). While the former envisions a two-step procedure – i.e. the forecasting is first performed at the top level and then, by disaggregating these data based on the historical percentage of each data point, within the whole group – in the latter each and every time series is first individually predicted and then all the forecasts are summed up. There is another approach which has gained widespread acceptance over the years, known as “middle-out”. It can be considered an extension of the top-down approach, since the forecast is first generated at two separate levels (upper and lower) and then combined in a proprietary manner to form a composite forecast. It is worth outlining how all of these methods can be considered sub-optimal insofar they neglect the correlation structure existing among the series belonging to the same level. Finally, a more recent approach known as optimal combination, envisions a two-step procedure where first the sequential and exhaustive forecast of each and every time series is independently performed and then – by optimally combining the predicted values obtained – are aggregated to achieve consistency across the hierarchical levels (reconciliation). Theoretically, cross-level coherency can be attained by means of Generalized Least Square (GLS) whose employment, however, turns out to be unfeasible due to the unidentifiability of the covariance matrix of the reconciliation errors (Wickramasuriya, S. L., Athanasopoulos, G., and Hyndman, R. J. (2019)). However, other methods, e.g. of the type OLS (Hyndman, R. J., Ahmed, R. A., Athanasopoulos, G., and Shang, H. L. (2011)) or WLS (Hyndman, R. J., Lee, A. J., and Wang, E. (2016)) can be used to circumvent this hurdle.

The method proposed in the present paper is of the type mixed, in the sense that exploits the approaches related to both forecasts reconciliation and forecasts combination. It is noted how mixed methods of this sort are not often encountered in literature. Such a situation might be due to the relatively recent introduction of reconciliation methods capable, unlike more traditional procedures, of accounting for the correlation structures among the series within a given hierarchical level, and thus able to deliver better performances. On the other hand, the need of a unified framework combining these two approaches has been recently brought up by Di Fonzo, T., and Girolimetto, D. (2020), which discussed an *ad hoc*, bi-dimensional (cross-sectional and temporal) procedure built upon a recent proposal by Wickramasuriya, S. L., Athanasopoulos, G., and Hyndman, R. J. (2019). Their method employs all the summation constraints arising in the cross-temporal hierarchic structure to reconciliate the base forecasts, using simple projections in a suitable linear space. On the other hand, the recent proposal by Spiliotis, E., Petropoulos, F., and Assimakopoulos, V. (2019) envisions a common framework where both forecast reconciliation and combination of forecasts generated by multiple models work together. Their method arises from the consideration that base forecasts should not be derived from a single method but a combination of methods. In the same direction goes the early work by Van Erven, T., and Cugliari, J. (2015), where both combination and reconciliation of the forecasts are applied in a two-step procedure, i.e. “first one comes up with the best possible forecasts for the time series without worrying about aggregation consistency and then a reconciliation procedure is used to make the forecasts aggregate consistent”. As it will be seen, the procedure illustrated in the present paper significantly differs from the above mentioned ones, in that different base forecasts are generated according to an arbitrary, pre-specified portfolio of statistical models, so that the combination exercise is performed on a set of already reconciled forecasts. Finally, while these authors focus on temporal aggregation this paper considers cross-sectional aggregations.

## 3. The framework

In this Section, an explanation of the framework within which the proposed method operates is given. In particular, the hierarchy structure of reference along with the employed reconciliation method are illustrated. The forecast combination part will be explained using two real-life examples of portfolios – which will be both used in the empirical Section – related to the statistical prediction models and the forecast combination techniques entertained.

### 3.1. Hierarchical cross-sectional reconciliation: the chosen method

This paper focuses on structures of the type summation constrained, in the sense that the underlying hierarchic structure of a given *m* – dimensional time series ***x***_*t*_, arises by summing up the bottom-level series into the higher ones. Figure 1 is an example of such a structure, under the condition that the constraints *x*_*t*_ = *x*_*a,t*_ + *x*_*b,t*_, *x*_*a,t*_ = *x*_*aa,t*_ + *x*_*ab,t*_ + *x*_*ac,t*_ and *x*_*b,t*_ = *x*_*ba,t*_ + *x*_*bb,t*_ are all satisfied.

**Fig. 1:**
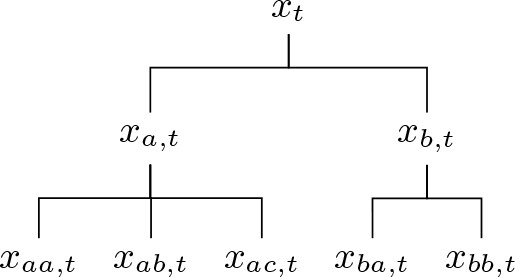
A two-level hierarchical structure

Formally, we have that the observed data ***x***_*t*_ – as well as their estimated future values, defined as ***x***_*h*_; *h* = 1, 2, …, *H*, with *H* the prediction horizon – lie in the summation-coherent subspace {𝒰}; *t* = 1, 2, …, *T* and ∀*h* = 1, 2, …, *H*. The prediction step subscript *h* has been omitted in Figure 1, for the sake of a better readability. In total, this hierarchy contains *m* = 8 time series, *n* = 5 of which are the lowest level time series, which therefore constitute the highest level of disaggregation. The observed series ***x***_*t*_ ∈ ℝ^*m*^ can be broken down as follows: 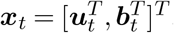, where 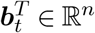 and 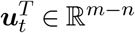 respectively contain the data pertaining to the bottom and upper series. Therefore, according this representation, the structure of Figure 1 (omitting the subscript *t*) can be broken down as follows: 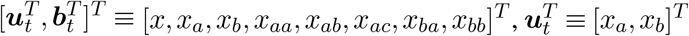 and 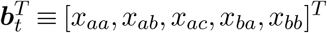. The hierarchic structure – satisfying ***x*** ⊂ {𝒰} – is induced by the summing matrix **𝒮** of dimension *m* × *n* such that ***x*** = **𝒮*b***_*t*_. Formally: ***x*** ⊂ {𝒰} ⟺ ***x***_*t*_ = **𝒮*b***_*t*_ (the symbol ⟺ replacing the locution “in and only if”). The **𝒮** matrix for the hierarchy in Figure 1 is as follows:

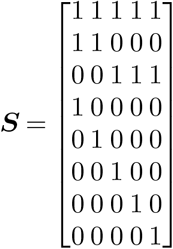

Using the symbols ∼ and ^ respectively to refer to the case of coherent and base (generally non coherent) forecasts, the reconciliated forecast *h*– step ahead can be expressed as proposed by Hyndman, R. J., Ahmed, R. A., Athanasopoulos, G., and Shang, H. L. (2011), i.e.

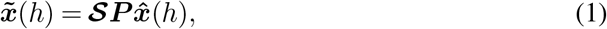

for some appropriately chosen matrix ***P* ∈** ℝ*m×n*. Assuming unbiased base forecasts, the best (in the sense of minimum sum of variances) linear unbiased revised forecasts are given by Equation 1 with

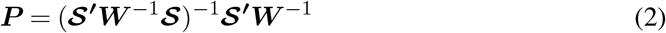

and thus (see Taieb, S. B., Taylor, J. W., andHyndman, R. J. (2017), Theorem 1)

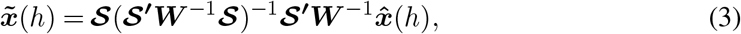

where **𝒮** is as above defined and 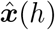 and 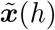; *h* = 1,2,…,*H* represent respectively the set of *H* predictions independently generated and the ones made coherent. Finally, ***W*** is the positive definite covariance matrix of the base forecast errors, i.e. 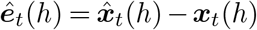, so that **W**(*h*) = 𝔼[*ê*_*t*_(*h*) − *ê*′_*t*_(*h*)]. As shown by Wickramasuriya, S. L., Athanasopoulos, G., and Hyndman, R. J. (2019), matrix ***W*** appears in the equation for the estimation of the error variance of the reconciled forecasts, i.e.

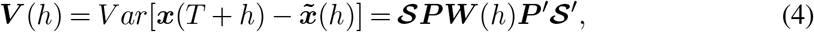

whose diagonal elements are the variances of the forecast errors. Their minimization can thus be performed in terms of the trace of ***V*** (*h*) and given by Equation 2 (therefore, this method is called Minimum Trace Estimator). Unfortunately, as proved by the same Authors, ***W*** is not identifiable, therefore, in the empirical section, the workaround proposed by them will be adopted. In essence, it is assumed ***W***_*h*_ = *k*(*h*)diag***Ŵ***1; *h* and assuming *k*(*h*) > 0 and denoting with ***W***_1_ the forecast errors covariance matrix estimated at horizon *h* = 1 – i.e. 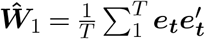– and with *K* is an unknown constant depending on the time horizon *h*.

### 3.2. The Forecast combination methods adopted

As already mentioned, the proposed method uses a set of combination methods, out of which the winner is selected according to a suitable loss function. In many empirical studies, it is shown how forecast combinations on average delivers better performances than methods based on a single forecasting statistical models. The theoretical validity of this approach is rooted in the assumption that the dimension of the sample sizes available in real-life applications are usually finite and, as a result, the correct specification of the “true” underlying data generation process is not attainable.

In what follows it is assumed ***x***_*t*_ to be the variable of interest and that ***f***_*t*_ = (*f*_1*t*_, *f*_2*t*_, …, *f*_*Nt*_)^′^ are the *N*, not perfectly collinear, available forecasts, whose combination is expressed as 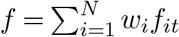, or, equivalently, 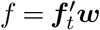, being *w*’s the combination weights.

The first method considered in this paper is of the type simple average. Despite its inherent simplicity (it ignores the correlation structure of the forecast errors) this method has been adopted given its ability, proved true in many cases, to “dominate more refined combination schemes aimed at estimating the theoretically optimal combination weights” (Atiya, A. F. (2020)). The simple average assigns equal weights to all predictors, i.e. 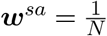 and thus the combined forecast is

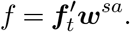

In the second method chosen, the forecast combination weights ***w***^*ols*^ = (*w*_1_, *w*_2_, …, *w*_*n*_), along with the intercept *b*, are computed using ordinary least squares (OLS) regression (Granger, C. W., and Ramanathan, R. (1984)), i.e.

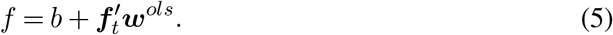

The third method applied – of the type Least Absolute Deviation (LAD) – is a modification of the OLS method, and it is expressed as in Equation 5, replacing the superscript *ols* with *lad*. Since the method of least squares assigns heavy weights on the error terms,the more robust estimator *LAD* – suggested by Gauss and Laplace – minimizes the absolute values and not the squared values of the error term. This features is particularly useful when the error term is generated by distributions having a infinite variance (fat tails) caused by outliers in the disturbance term.

Finally, a modification of the method proposed by Newbold, P., and Granger, C. W. (1974), built upon an earlier methodology of Bates, J. M., and Granger, C. W. (1969), is our fourth approach. Let be the positive definite matrix of the mean squared prediction errors (MSPE) of ***f***_*t*_ and ***g*** is an *N* × 1 vector of (1, 1, …, 1)′ their method relies on a constrained minimization of the MSPE under the normalizing condition ***g***′***w*** = 1. The resulting combination of weights is

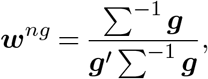

so that the combined forecast is

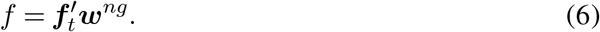

However, unlike the original method, the variant employed here follows the proposal by Hsiao, C., and Wan, S. K. (2014), which does not impose the prior restriction that the matrix ∑ is diagonal.

### 3.3. The entertained statistical models

Before delving into the proposed method, a quick presentation of the forecasting methods employed in the empirical section is in order. The first two statistical models considered are of the type ARIMA (Auto Regressive Fractional Moving Average) (Box, G. E., Jenkins, G. M., and Reinsel, G. (1970)) and *ARF IMA* (Auto Regressive Fractional Moving Average) (Granger, C. W., and Ramanathan, R. (1984) and Granger, C. W., and Joyeux, R. (1980)). Being the latter a generalization of the former, the two models will be presented conjointly.

*ARF IMA* (Auto Regressive Fractional Moving Average) models are useful in circumstances where the underlying stochastic process exhibits hyperbolic decay patterns in their estimated autocorrelation function. ARFIMA-type processes are usually expressed as follows:

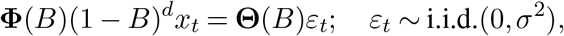

where *d* is a parameter – assumed to take non-integer values in the difference operator (1 − *B*)^*d*^, with *B* identifying the backward operator, that is *B*^*k*^*x*_*t*_ = *x*_*t*−*k*_. The fractional differencing operator is defined by the binomial expansion 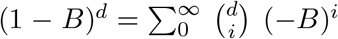. The process is stationary and invertible if the roots of the autoregressive polynomial of order *p*, **Φ**(*B*) = 1 − *ϕ*_1_*B* − *ϕ*_2_*B*^2^, …, −*ϕ*_*p*_*B*^*p*^, and the order *q* moving-average part, **Θ**(*B*) = 1 + *θ*_1_*B* + *θ*_2_*B*^2^ + … + *θ*_*q*_*B*^*q*^, lie outside the unit circle with |*d*| < 0.5.

ARFIMA models generalize the ARIMA(p,d,q) representation where the parameter *d* is constrained to integer values. This type of model has been designed to capture approximately parabolic decay patterns of the empirical autocorrelation function. As such, they are suitable to model persistence structures embedded in the underlying stochastic process of the type short-memory.

Theta method – the third forecasting model considered – is a powerful class of models which have been proposed by Assimakopoulos, V., and Nikolopoulos, K. (2000). Define with the symbol ∇ the difference operator – i.e. ∇*x*_*t*_ = *x*_*t*_ − *x*_*t* − 1_, *x*_*t*_ being the original time series – this method is the solution of the equation

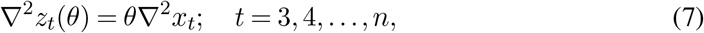

with *z*_*t*_(*θ*)’s analytical solution reading as following: *z*_*t*_(*θ*)*θx*_*t*_ + (1 − *θ*)(*A*_*n*_ + *B*_*n*_*t*); *t* = 1, 2, …, *n*, where *A*_*n*_ and *B*_*n*_ are the minimum square coefficient of a linear regression equation of the series *x*_*t*_ against **1**_*n*_, i.e. the vector of ones of length *n*. These terms are given by 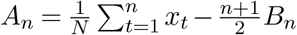 and 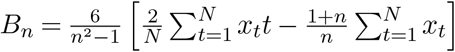. Finally, the initial values *z*_1_ and *z*_2_ in Equation 7 are estimated by minimization of 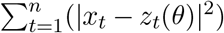.

The fourth and last model employed is of the type exponential smoothing, proposed in 1944 by Robert G. Brown, a US Navy operations research analyst (Gardner Jr, E. S. (2006)). Specifically, two schemes have been employed here: Additive Holt Error Model (*AEM*) and multiplicative Holt Error Model (*MEM*) (as it will be seen, the procedure automatically will select the “best” one). As for the *AEM*, let 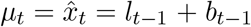 be the one-step ahead forecast of the observed time series *x*_*t*_ generated by the forecasting equation *x*_*t*_ = *l*_*t*−1_ + *b*_*t*−1_ + *ε*_*t*_, being *l*_*t*_ a measure of the level of the series, *b*_*t*_ an estimate of the slope (or growth) at time t and *ε*_*t*_ = *x*_*t*_ − *µ*_*t*_ the one-step-ahead forecast error, referred to the time *t*. The level and slope equations for *AEM* are respectively represented as

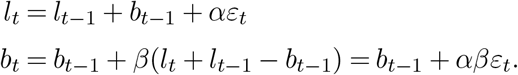

By re-expressing the error term *ε*_*t*_ as 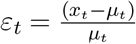 (relative errors), the forecast, level and slope equations for the *MAM* model are as follows:

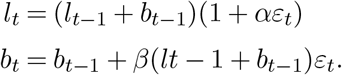

In the above two sets of equations *α* and *β* are the model parameters to be estimated.

## 4. The proposed method

Let us indicate with the symbols **ℛ** and |· | respectively a suitable reconciliation method and the cardinality function (assuming the number of elements in a given set to be finite, |· | simply returns the number of the elements belonging to that set). Let the symbol **ncol** identify the function which, applied to a given matrix, returns its number of columns and **ℳ** ≡ {*µ*_1_, *µ*_2_, …, *µ*_*M*_} and **𝒟** ≡ {*δ*_1_, *δ*_2_, …, *δ*_*D*_} respectively the set of | **ℳ** |= *M* prediction models and the set |**𝒟**| = *D* of forecast combination methods entertained, both arbitrarily chosen. Once applied to the time series of interest *x*_*t*_; *t* = 1, 2, …, *T*, each model {*µ*_*j*_ ∈ **ℳ**; *j* = 1, 2, … *M* generates a set, called **ℱ**^*H*^, made up with *M H* –step ahead predictions, i.e.: {**ℱ**^*H*^(*µ*_*j*_); *j* = 1, 2, … *M*}. Each of the elements of this set is a base forecasts, in the sense that it is generated by individually applying *a* given statistical model *µ*_*j*_ to the observed time series without any attempt of reconciliation.

Each of these *M* elements in ***ℱ*** (the *M* forecast vectors) is individually reconciled through the reconciliation procedure **ℝ**, i.e. {**ℝ** (***ℱ*** (*µ*_*j*_); *j* = 1, 2, … *M*)} (the superscript *h* is omitted for brevity). At this point, the resulting set ***𝒫***(*µ*_*j*_); *j* = 1, 2, … *M* of *M* model–dependent reconciled forecasts (first optimization) is optimally combined by applying each method in the set ***𝒟*** to any possible combination (without repetition) of order {*k* = 1, 2, …, *M*} to the set ***𝒫*** (second optimization). The resulting set ***Ƶ*** – with cardinality 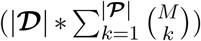 – contains all the possible combinations – ∀*k*-order – of the model-dependent reconciled forecasts. The third optimization step is carried out by applying to ***Ƶ*** a suitable loss function, here denoted with the symbol ***ℒ***(·). The optimal vector of forecasts is thus the element ***z***\* ∈ *Ƶ* minimizing this function, i.e. ***z***\* = min ***ℒ***(***Ƶ***). This optimality condition is expressed as

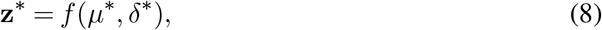

being the arguments of *f* respectively the “best” forecasting model and forecast combination technique. This last step, by ruling out the less performing combination method(s), has been introduced in order to reduce the overall uncertainty level of the analysis. In fact, suppose that the original set ***𝒟*** reduces to ***𝒟***′ – being clearly |***𝒟***′| < |***𝒟***| – the additional amounts of undesired fluctuations and noise – which one can reasonably expect as a consequence of the employment of one (more) under-performing combination method(s) – are avoided. Finally, the model bias *β*\* is empirically estimated using the in–sample residuals generated by employing the winners techniques *µ*\* and *δ*\*, according to an optimal choice made on a set of suitable central tendency functions (fourth optimization).

For the sake of clarity, a more schematic description of the method is given below, in the form of algorithm presented in a step-by-step fashion.

**Figure 1.**
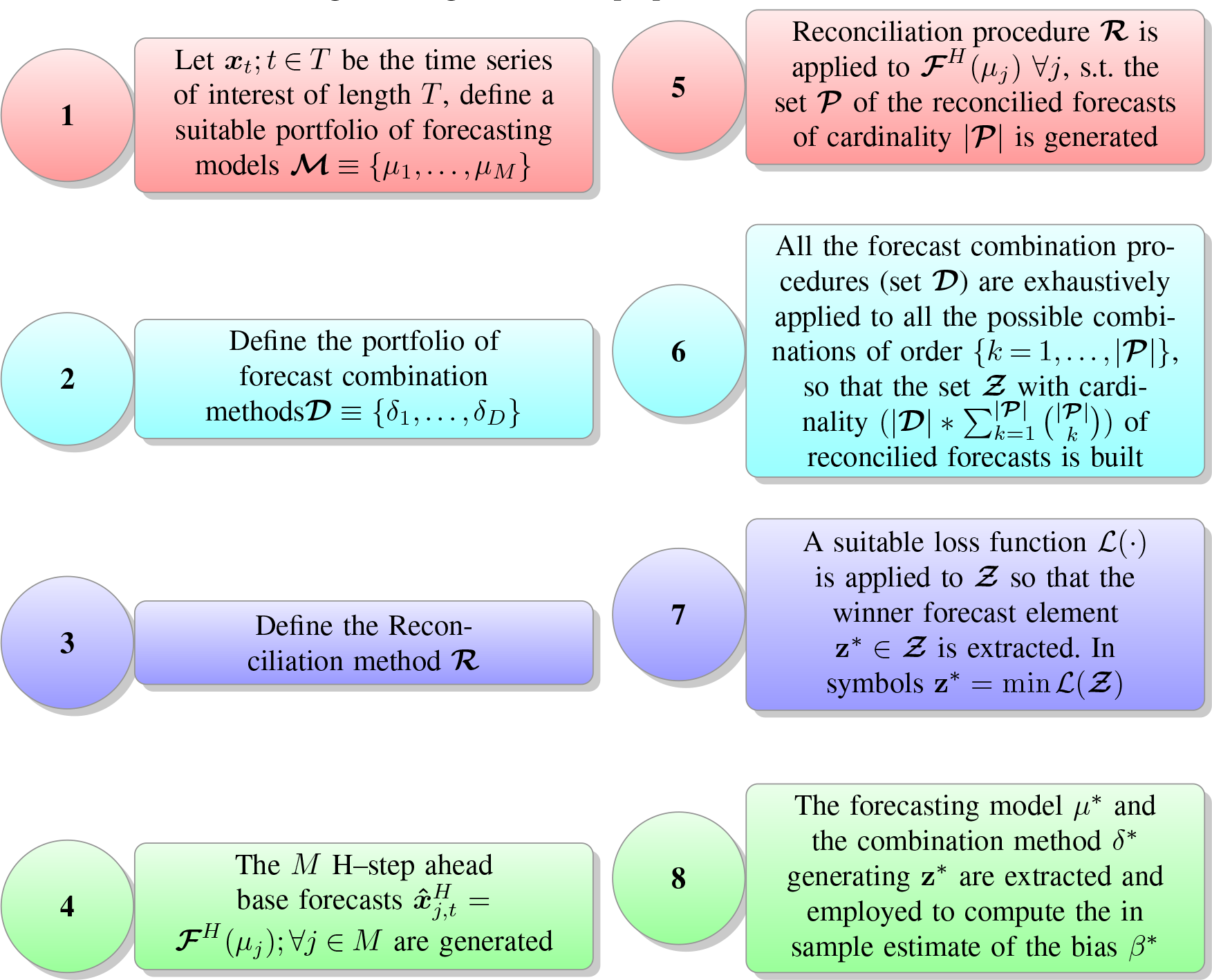
Algorithm of the proposed method.

The novelty of the proposed method is basically captured by steps 5–7 and 8. For the sake of a more operational comprehension, step 5–7 are now discussed using the matrix notation whereas step 8 will be detailed in Subsection 4.1. Step 5 indicates that once a number *M* of different forecasts, generated by *M* models, become available for each level of the hierarchy, they are reconcilied one at a time through **ℛ**. In practice, the reconciliation function **ℛ**, applied to the given hierarchic structure, generates a sequence of *H* × 1 vectors of reconciled predictions 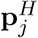, as below schematized.

**Figure 2.**
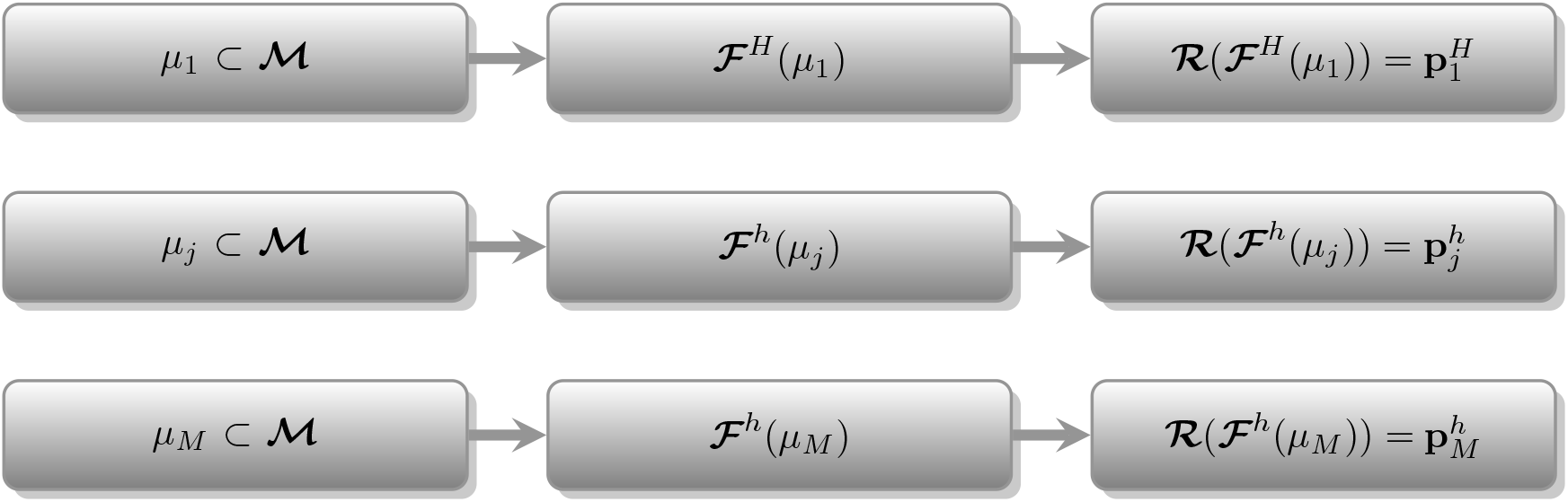
Illustration of the procedure to obtain the vectors of reconcilied forecasts.

Each vector **p**_*H,j*_ can be seen as the column of a matrix, say **P**_*H,M*_, containing all the *M* model-dependent reconcilied forecasts. In the following step 6, these *M* (column)-vectors of predictions are combined according to a number *D* of different combination methods (*δ*_1_, …, *δ*_*D*_). In essence, they are sequencially and exhaustively applied to each of the possible combinations of order {*k* = 1, …, ncol(***P***)} of the column vectors of **P**_*H,M*_. Defining the combination (without repetition) function with *C*_*k*_ and setting, for instance, the combination order *k* = *k*_0_ < *M*, the submatrix 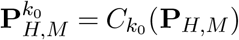 stores all the 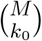 combinations of the forecasts 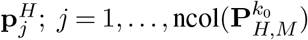, called 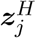, as illustrated in the Figure below.

**Figure 3.**
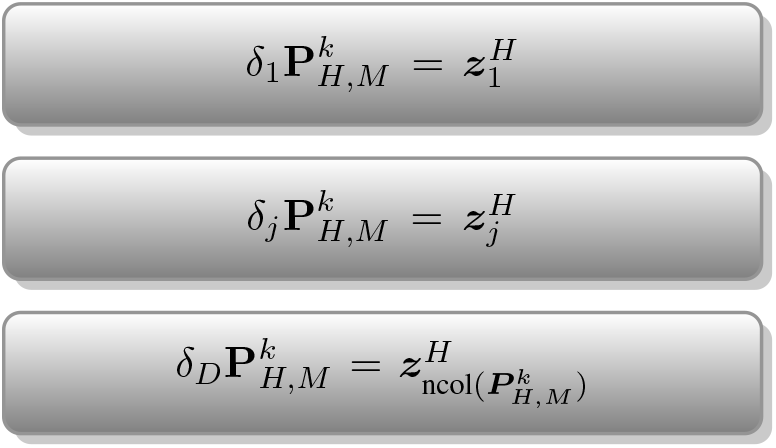
Illustration of the procedure to obtain the vectors of model dependent reconcilied forecasts.

By looping over all of the combination orders *k* = 1, …, ncol(**P**_*H,M*_), the matrix ***Z*** containing all the possible combination of the *M* model dependent reconcilied forecasts is obtained. This matrix is called ***Z*** and has dimensions

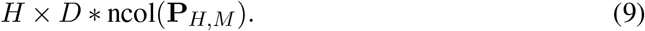

Step 7 translate into simply applying a suitable loss function to ***Z*** (column-wise), until the final vector of predictions ***z***\*, verifying the minimum condition min *ℒ*(**Z**), is extracted.

### 4.1. The bias correction procedure (step 8)

It is well known that a perfectly unbiased forecast is a condition not frequently met in many real-life applications. Unfortunately, the unbiasedness of the forecasts reconciliation method chosen (Equation 2), depends on the unbiasedness of each and every base forecasts, as proved by Wickramasuriya, S. L., Athanasopoulos, G., and Hyndman, R. J. (2019). The proposed method can alleviate this problem as it is designed to generate a “big” competition set (***Ƶ***), made up with more “balanced” forecasts (thanks to the forecast combinations techniques applied) and thus more likely to perform better than methods generating fewer or just one forecast vector.

The bias correction of the forecast values stored in the vector **z**\* – obtained in step 7 of Figure 1 – is performed using an improved version of the simple, yet effective, procedure discussed in Spiliotis, E., Petropoulos, F., and Assimakopoulos, V. (2019). In more details, the adopted method translates into a six–step iterative procedure, designed to empirically estimate a set of in-sample tentative biases *{****β*** ≡ *β*_1_, *β*_2_, …, *β*_*B*_*}*, each of them obtained according to a predefined, arbitrary set of suitable central tendency functions *{a*_1_, *a*_2_, …, *a*_*A*_ ⊂ ***A****}*, being |***β***| = |***𝒜***| (or, equivalently, *B* = *A*).

Recalling that ***x***_*t*_ is the observed time series, let us denote with 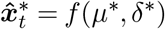 its one-step-ahead predictions – obtained according to the optimal forecasting model and prediction combination method (see Equation 8) – and with ***ε***_***t***_|*β*_*j*_ the vector of residuals between these two series conditional to a given central tendency function, i.e. 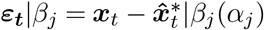 In what follows, the term *α*_*j*_ is omitted as it is understood the dependency relationship between bias and central tendency function, i.e. bias = *β*_*j*_(*α*_*j*_). The set ***β*** is thus generated by iteratively and exhaustively applying each function in **𝒜** to the bias adjusting equations which, once expressed in terms of residuals, read as follows:

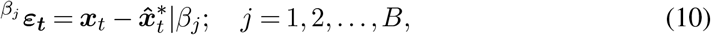

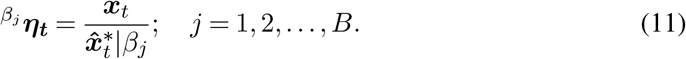

Equations 10 – 11 differ only for that the one-step ahead predictions are respectively adjusted additively and multiplicatively. Finally, by applying a suitable loss function, () to each of the vectors 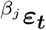 and 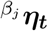, the optimal bias estimation is its minimzer, i.e.

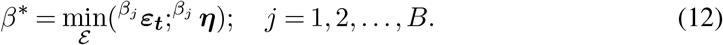

Once the final bias is computed, it can be readily applied in a forward looking fashion, i.e.

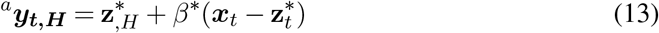

or

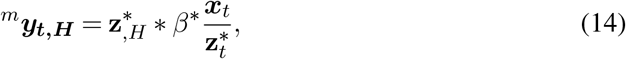

according to whether the winner central tendency function is applied in an additive (Equation 13) or multiplicative fashion (Equation 14). The generilzed notations for the first term in Equations 4.1 and 4.1 is

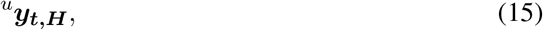

which represent the final predictor. In such an approach, the future and the past are assumed to be affected by the same amount of bias. Such an assumption, under stationaity of the observed time series and a “sufficient” sample size, might not be considered unreasonable.

In the case of {***β*** ≡ *β*_1_} with *β*_1_ the mean function, Equations 13–14 are respectively as follows:

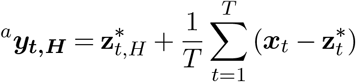

or

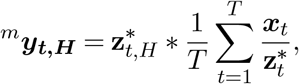

and thus equivalent (saved for the notation) to the procedure discussed in Spiliotis, E., Petropoulos, F., and Assimakopoulos, V. (2019) (Equations 3–4), page 21.

In the empirical application of Section 6, the central tendency functions applied are: mean, median and root mean square, respectively denoted by the lowercase Greek letters *π, τ* and *ξ*. Their mathematical representations is as follows:

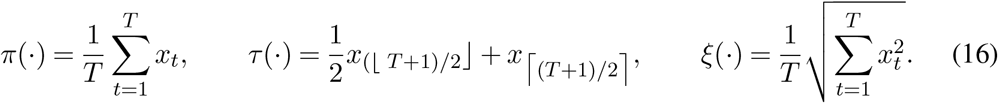

In the case of the median (*τ* (·)) *x* is an ordered list of T values, and the symbols ⌊·⌋ ⌈·⌉ denote the floor and ceiling functions, respectively.

## 5. Justification of the method

The effectiveness of the proposed method is, in general, conditioned to the choices of the statistical prediction models and the forecast combination techniques included in the sets **ℳ** and **𝒟**, as well as to the selection of the most suitable central tendency functions (the set **𝒜**). A careful building of those sets (our multidimensional search space), is a prerequisite for the proposed method to properly perform. Its final dimensions are as in Expression 9 plus twice the number of central density functions considered, used in both additive and multiplicative fashion, i.e.

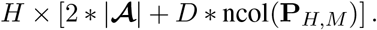

The point of strength of the method is thus related to the availability of a potentially large number of multiple choices, all of them derived in a multiple combination set-up (in terms of statistical prediction model, forecast combination and bias correction), so that the selected forecast vector are the minimizer of a bi-dimensional loss function (ℒ,*ε*). In addition, the method is very flexible, as it can work with all the methods deemed suitable for the problem at hand, being the only limit the computational time. This is certainly an issue, which, however, can be easily circumvented thanks to the structure of the method itself, which is naturally prone to be parallelized.

According to the bias–variance decomposition approach, the mean square error (*MSE*) can be decomposed into a bias *β* and a variance (*V*) terms, i.e.

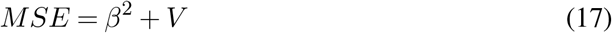

In what follows, the advantages related to the proposed method will be illustrated in terms of Equation 17. Firstly, it is noted how, in general, simpler models tend to produce large biases and small variances whereas complex models behave in the opposite way. The proposed method is designed to overcome such an issue not only because it can employ a combination of several models with different levels of complexity but also because it selects the “best” combination technique (stored in the set ***𝒟***) according to the data set under investigation. This last feature is clearly a plus, since there is not such combination techniques able to perform optimally in any circumstances. For example, Palm, F. C., and Zellner, A. (1992) found that there are cases where a simple average combination may be more robust than weighted average combinations. Therefore, by iteratively testing many different techniques, one is more likely to find the most suitable (if not the “optimal”) one.

Bias-wise, the advantages of this method are related to its self-balancing and self-adjusting features, the former being induced by the bias compensation phenomenon, more likely to occur in a multi-model set up, whereas the latter relies on the bias correction procedure, given in Section 4.1. In particular, its effectiveness in bias reduction is related to the fact that the self-adjustment procedure uses a vector of forecast which, by design, has already been controlled for bias. To see this, let us express the generic forecast combination 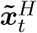 as

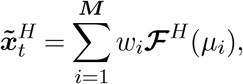

with 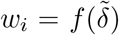 the combination weights generated by the combination method 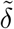 and ***ℱ***^*H*^ (*µ*_*j*_); *j* = 1, …, *M* are base forecasts generated by the *M* statistical models entertained (see Figure 1 step 1 and 4). Assuming 0 ≤ *w*_*j*_ ≤ 1, 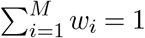 and the vector of “future” observations of length *H* to be known, the total amount of bias of the combined forecast is given by

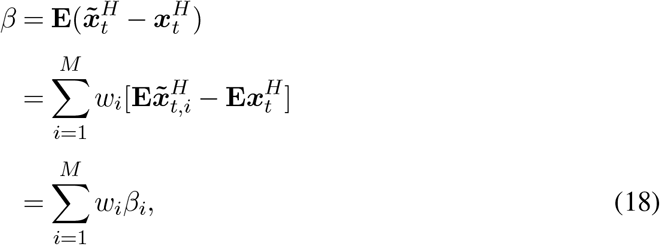

where the subscript *i* is used to refer to a specific model, in terms of generated bias (*β*_*i*_) and forecast 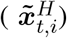. The right term of Equation 18 shows that the bias of the forecast combination is the weighted average of the biases of the base forecasts and thus, provided that their magnitude is comparable, one can reasonably expect an overall bias reduction due to cancellation effects. Such a phenomenon is not rare, since – in general – it is not common for all the biases to show the same sign. Since the bias-correcting method – discussed in Section 4.1 – is applied on an already optimally combined vector of forecasts 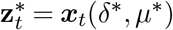, a less pronounced bias can be expected in the final predictions, given by

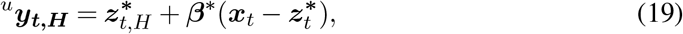

where *β*\* is as in Equation 12.

The proposed method can also help keep low the variance of the forecasts in an amount inversely proportional to the correlation coefficients computed between the competing forecasts and proportional to the reduction in the standard errors induced by the reconciliation procedure adopted ℝ. To see this, denoting with 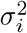 the variance of the individual forecast *i* and with *γ*_*i,j*_ the correlation coefficient computed on a generic pair (*i, j*) of forecasts, we use the following inequality (derived in Atiya, A. F. (2020)), i.e.

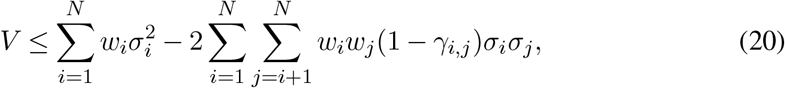

where *V* is the variance of the combination of forecasts. Inequality 20 states that *V* tends to be considerably less than the average of the individual forecasts in an amount depending on 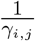, meaning that less correlated forecasts are beneficial in terms of variance reduction. Such a situation is more likely to occur in procedures which, as the one proposed, grant a “sufficient” number of predictions. However, there is another point in favor of the proposed method on this matter. In fact, recalling that the different combinations of the forecasts are performed on already reconciled vectors of predictions 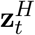 – according to the adopted procedure ℛ– by virtue of Equation 4 their variances obey to the following inequality:

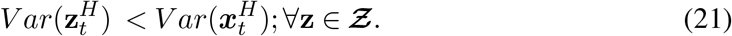

Therefore, the overall level of variance in Equation 20 decreases of an amount inversely proportional to *V ar*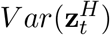.

## 6. Empirical study

In this section the goodness of the proposed method will be evaluated using the official time series related to the number of people tested positive to the SARS-CoV-2 in each of the Italian regions, between February 24^*th*^ 2020 and October 7^*th*^ 2020. The whole data set – issued by the Italian National Institute of Health – are publicly and freely available at the web address *https://github.com/pcm-dpc/COVID-19/tree/master/dati-regioni*. The data, sampled at a daily frequency, are stored in a matrix called **𝒪** (see Table 1) of dimension 227 *×* 21, where 21 are the Italian regions. From a strictly administrative point of view, the number of the Italian regions amounts to 20, however, for one of them, called Trentino Alto Adige, the data are split according to its two main towns: Trento and Bolzano. As reported in the same Table 1, the proposed procedure is trained on a portion of the data matrix, called **𝒪**^*train*^, of dimensions 197 *×* 21 and time span February 24^*th*^ – September 7^*th*^, whereas the test part is carried out on a set, called **𝒪**^*test*^, whose dimensions are *H* = 30 *×* 21 (the time span is from September 8^*th*^ to October 7^*th*^). Finally, 30 days ahead “real-life” estimates – in the sense that they are related to future values which are unknown at the time of their computations – for the time window October 8^*th*^ – November 7^*th*^ will be stored in the matrix **𝒪**^*fore*^. Since the proposed procedure combines a number of models (*µ*’s), combination methods (*δ*’s) and central tendency functions (*ε*^*t*^*s*), for each of those, conventional symbols are respectively given in Tables 2, 3 and 4. Consistently with the convention introduced in Equation 15, in Table 4 the superscript *u* is used to indicate the type of bias considered, i.e. additive (*u* = *a*) or multiplicative (*u* = *m*). Finally, to efficiently keep track of the outcomes of the method, in Table 5 the whole set of model combinations employed – respectively of class *k* = 4, 3, 2 – are provided. Since we have four different combination methods, each of the 11 model combination (reported in Table 5) are performed four times, which yields a total of 44 method-dependent combinations. One of them, for example, is the forecasts combination generated by combining an ARFIMA and an ETS models using the method Ordinary Least Square. This information is conveniently conveyed by the symbol *OLS–AE*.

**Table 1.** The employed data set and its portions defined according to the different purposes served

| Symbol | Start date | End Date | Sample size |
| --- | --- | --- | --- |
| $\mathcal{O}$ | February 24<sup>th</sup> | October 7<sup>th</sup> | 227 |
| $\mathcal{O}^{train}$ | February 24<sup>th</sup> | September 7<sup>th</sup> | 197 |
| $\mathcal{O}^{test}$ | September 8<sup>th</sup> | October 7<sup>th</sup> | 30 |
| $\mathcal{O}^{fore}$ | October 8<sup>th</sup> | November 7<sup>th</sup> | 30 |

**Table 2.** Symbols employed to identify the statistical models

| Model ($\mu$) | Symbol |
| --- | --- |
| ARFIMA | A |
| ETS | E |
| THETA | T |
| ARIMA | B |

**Table 3.** Symbols employed to identify the central tendency functions

| Method ($\delta$) | Symbol |
| --- | --- |
| Ordinary Least Square | OLS |
| Least Absolute Deviation | LAD |
| Newbold and Granger | NG |
| Simple Average | SA |

**Table 4.** Symbols employed to identify the statistical models

| Central tendency function | Symbol |
| --- | --- |
| Mean | ${}^u\pi$ |
| Median | ${}^u\tau$ |
| RMS | ${}^u\xi$ |

**Table 5.** Combinations of models of class K = 4, 3, 2 attempted for each of the four model-dependent reconcilied forecasts

| Number | $K$ | Model combination |
| --- | --- | --- |
| 1 | 4 | A-E-T-B |
| 2 | 3 | A-B-E |
| 3 |  | A-B-T |
| 4 |  | A-E-T |
| 5 |  | B-E-T |
| 6 | 2 | A-B |
| 7 |  | A-E |
| 8 |  | A-T |
| 9 |  | B-E |
| 10 |  | B-T |
| 11 |  | E-T |

Recalling that with ^*u*^***y***_***t***,***H***_ the final predictions yielded by the proposed method are denoted (see Equation 15), the loss function employed (ℒ) is the Root Mean Square Error (RMSE), given by 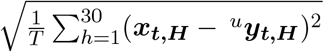. The same function is adopted in–sample to select the best 3–tuple (*µ*^*^, *δ*^*^, *ε*^*^), i.e 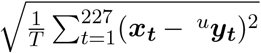 and to evaluate the method’s performance in the test set **𝒪**^*test*^. Finally, the out of sample estimate of the bias, in the sequel denoted by the symbol *β*^*out*^, has been computed on the set **𝒪**^*test*^, using the formula 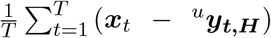.

### 6.1. Performances of the method

The performance of the method are summarized in Appendix A and in Table 6. In particular, in Appendix A the observations belonging to the test set **𝒪**^*test*^ (black line) and the related estimates ^*u*^***y***_***t***,***H***_ (red line) are depicted. The first three columns of Table 6 respectively indicate the name of the Italian regions, the winner combination (*µ*^*^, *δ*^*^) and the related RMSE values (ℒ^*^). The best central tendency function (*ε*^*^), the selected bias corrector(*β*^*^) and the estimated out-of-sample bias (*β*^*out*^) are given in columns four, five, six. In the last two columns the *RMSE* values (ℒ) relative to each of the forecasting models (reported in Table 2) taken separately, are recorded. The accuracy of the proposed method seems to be very good as, in almost all the cases, the winner combinations deliver better predictions than all the statistical models singularly considered and, in many cases, outperform them. This is the case, for example, of the Campania and Calabria regions. Here (see Table 6), the recorded *RMSE* is respectively equal to 34.7 and 63.5, far below the values obtained using the best statistical models, i.e. *Theta* and *ETS*, which respectively scored ℒ = 143.6 and 265.1. In addition, the proposed procedure shows always from very low to negligible in-sample amounts of bias. The most selected central tendency functions is ^*a*^*τ* and ^*a*^*π*, whereas it is noted that the *RMS* function (^*u*^*ξ*) has never been chosen. Regarding the out-of-sample bias, if on one hand, as expected, it is always *β*^*out*^ *> β*^*^, on the other hand it can be said that the magnitude of the values assumed by *β*^*out*^ can be deemed acceptable. This is especially true if one considers the length of the prediction window (*H* = 30 days) compared with the available data set. In particular, six regions (Valle d’Aosta, Trento, Molise, Lazio, Abruzzo, Calabria) show interesting values for the out of sample bias, being *β*^*out*^ < 10. The worse performances of the method refers to the region of Sardegna, where the consistent underestimations of the true values lead to a recorded bias of around 504. This fact can be explained by looking at the irregular, bumpy shape of this time series, reported in Appendix 2 (Sardegna), which might have introduced distortions in the model estimators. In Appendix B, the graphical results of a pure out-of-sample application of each of the winner combinations are reported. In more details, the region-specific winner combinations (*µ*^*^, *δ*^*^) are applied to the whole set **𝒪**, so that the *H* = 30 days–ahead resulting forecasts – stored in the set **𝒪**^*fore*^ – are the pure forecasts for the period October 8^*th*^ – November 7^*th*^. In Appendix B, the regional time series in **𝒪** (true observations) are plotted in black whereas the predictions are in red color. The analysis of these Figures suggest a slower acceleration in the growth of positive cases in some of the north regions (e.g. Lombardia, Trento, Liguria) whereas the center and south regions might face a strong increase of positives. This seems likely to happen in the Campania, Basilicata and Molise regions. The number of positive for Italy, predicted for the end of the pure forecast period (November 7^*th*^), is about of 115,854.

**Table 6:** Performances of the method for each of the Italian regions. Outcomes of the winner models and of the single statistical models. Refer to the text for details

| Region | Winner combination | $\mathcal{L}^*$ | $\mathcal{E}^*$ | $\beta^*$ | $\beta^{out}$ | Single models | $\mathcal{L}$ |
| --- | --- | --- | --- | --- | --- | --- | --- |
| Piemonte | $LAD - BET$ | 135.6 | $a_{\tau}$ | $\approx 0$ | 54.09 | ARFIMA<br>ETS<br>$\theta$<br>ARIMA | 1050.8<br>1065.8<br>1271.8<br>683.6 |
| Val d'Aosta | $SA - BT$ | 8.6 | $a_{\tau}$ | $\approx 0$ | -2.25 | ARFIMA<br>ETS<br>$\theta$<br>ARIMA | 60.7<br>177.5<br>30.5<br>29.5 |
| Lombardia | $SA - ET$ | 172.6 | $m_{\tau}$ | 1.0 | 70.83 | ARFIMA<br>ETS<br>$\theta$<br>ARIMA | 3184.0<br>911.7<br>966.1<br>1713.8 |
| Bolzano | $OLS - ET$ | 223.2 | $a_{\pi}$ | $\approx 0$ | 190.9 | ARFIMA<br>ETS<br>$\theta$<br>ARIMA | 468.9<br>227.6<br>271.2<br>248.8 |
| Trento | $NG - BE$ | 29.4 | $a_{\tau}$ | -0.13 | 8.95 | ARFIMA<br>ETS<br>$\theta$<br>ARIMA | 113.7<br>30.0<br>238.2<br>211.5 |
| Veneto | $NG - BE$ | 58.5 | $a_{\pi}$ | 0.55 | 49.01 | ARFIMA<br>ETS<br>$\theta$<br>ARIMA | 620.1<br>95.5<br>259.9<br>49.6 |
| Friuli Venezia Giulia | $SA - BE$ | 65.9 | $a_{\pi}$ | 0.70 | 18.08 | ARFIMA<br>ETS<br>$\theta$<br>ARIMA | 475.4<br>140.6<br>763.5<br>155.6 |
| Liguria | $LAD - ET$ | 124.2 | $a_{\tau}$ | $\approx 0$ | -28.13 | ARFIMA<br>ETS<br>$\theta$<br>ARIMA | 4123.8<br>126.8<br>1057.7<br>241.9 |
| Emilia Romagna | $LAD - BT$ | 296.7 | $a_{\tau}$ | $\approx 0$ | -160.01 | ARFIMA<br>ETS<br>$\theta$<br>ARIMA | 2203.4<br>343.5<br>724.5<br>274.9 |
| Toscana | $LAD - AB$ | 259.6 | $a_{\tau}$ | $\approx 0$ | 149.8 | ARFIMA<br>ETS<br>$\theta$<br>ARIMA | 3417.7<br>1202.4<br>2485.9<br>310.0 |
| Umbria | $OLS - AB$ | 133.3 | $a_{\pi}$ | $\approx 0$ | 104.54 | ARFIMA<br>ETS<br>$\theta$<br>ARIMA | 701.9<br>254.5<br>344.4<br>168.1 |

| Region | model | $\mathcal{L}^*$ | $\mathcal{E}^*$ | $\beta^*$ | $\alpha^*$ | Single models | $\mathcal{L}$ |
| --- | --- | --- | --- | --- | --- | --- | --- |
| Marche | $SA - BET$ | 211.0 | $m_{\tau}$ | 1.01 | 136.65 | ARFIMA<br>ETS<br>$\theta$<br>ARIMA | 2191.6<br>320<br>1185.7<br>355.5 |
| Lazio | $SA - ET$ | 45.9 | $a_{\tau}$ | 0.13 | 7.43 | ARFIMA<br>ETS<br>$\theta$<br>ARIMA | 522.2<br>153.5<br>180.7<br>64.7 |
| Abruzzo | $LAD - BET$ | 40.0 | $a_{\tau}$ | $\approx 0$ | 6.97 | ARFIMA<br>ETS<br>$\theta$<br>ARIMA | 740.2<br>70.3<br>294.6<br>40.7 |
| Molise | $SA - AB$ | 39.2 | $a_{\pi}$ | -.05 | -5.8 | ARFIMA<br>ETS<br>$\theta$<br>ARIMA | 250.9<br>1229.0<br>147.0<br>265.1 |
| Campania | $SA - BT$ | 34.7 | $a_{\tau}$ | .05 | -30.52 | ARFIMA<br>ETS<br>$\theta$<br>ARIMA | 480.7<br>359.8<br>143.6<br>201.3 |
| Puglia | $LAD - BET$ | 419.4 | $a_{\tau}$ | $\approx 0$ | -249.58 | ARFIMA<br>ETS<br>$\theta$<br>ARIMA | 1991.5<br>680.6<br>2507.5<br>674.1 |
| Basilicata | $LAD - AB$ | 75.2 | $a_{\tau}$ | $\approx 0$ | -54.68 | ARFIMA<br>ETS<br>$\theta$<br>ARIMA | 117.5<br>1453.5<br>37.0<br>417.7 |
| Calabria | $OLS - AETB$ | 63.5 | $a_{\pi}$ | $\approx 0$ | 4.13 | ARFIMA<br>ETS<br>$\theta$<br>ARIMA | 2124.9<br>265.1<br>1109.9<br>302.1 |
| Sicilia | $SA - BET$ | 70.7 | $m_{\pi}$ | 0.39 | 57.68 | ARFIMA<br>ETS<br>$\theta$<br>ARIMA | 1436.8<br>333.0<br>715.7<br>195.2 |
| Sardegna | $NG - AB$ | 581.9 | $m_{\pi}$ | -0.05 | 504.75 | ARFIMA<br>ETS<br>$\theta$<br>ARIMA | 983.7<br>998.9<br>1204.6<br>605.4 |

## 7. Conclusions and future directions

The present paper provides sufficient evidences that reconciliation serves the double purpose of generating coherent forecast with improved accuracy, under a four dimensional optimization constraint. In fact, the proposed method is designed to handle the increased amount of uncertainty surrounding the forecasting, as one carries out the prediction exercise at a progressively more disaggregated levels.

## Data Availability

Data availability. The data that support the findings of this study are openly available in the section COVID-19/dati-regioni/ at the https://github.com/pcm-dpc/COVID-19/tree/master/dati-regioni.

## 8. Data availability

The data that support the findings of this study are openly available in the section “COVID-19/dati-regioni/” at the https://github.com/pcm-dpc/COVID-19/tree/master/dati-regioni.

## 9. Disclaimer

The views and opinions expressed in this article are those of the author and do not necessarily reflect the official policy or position of the Italian National Institute of Statistics.

## 10. Appendix A

**Fig. 5:**
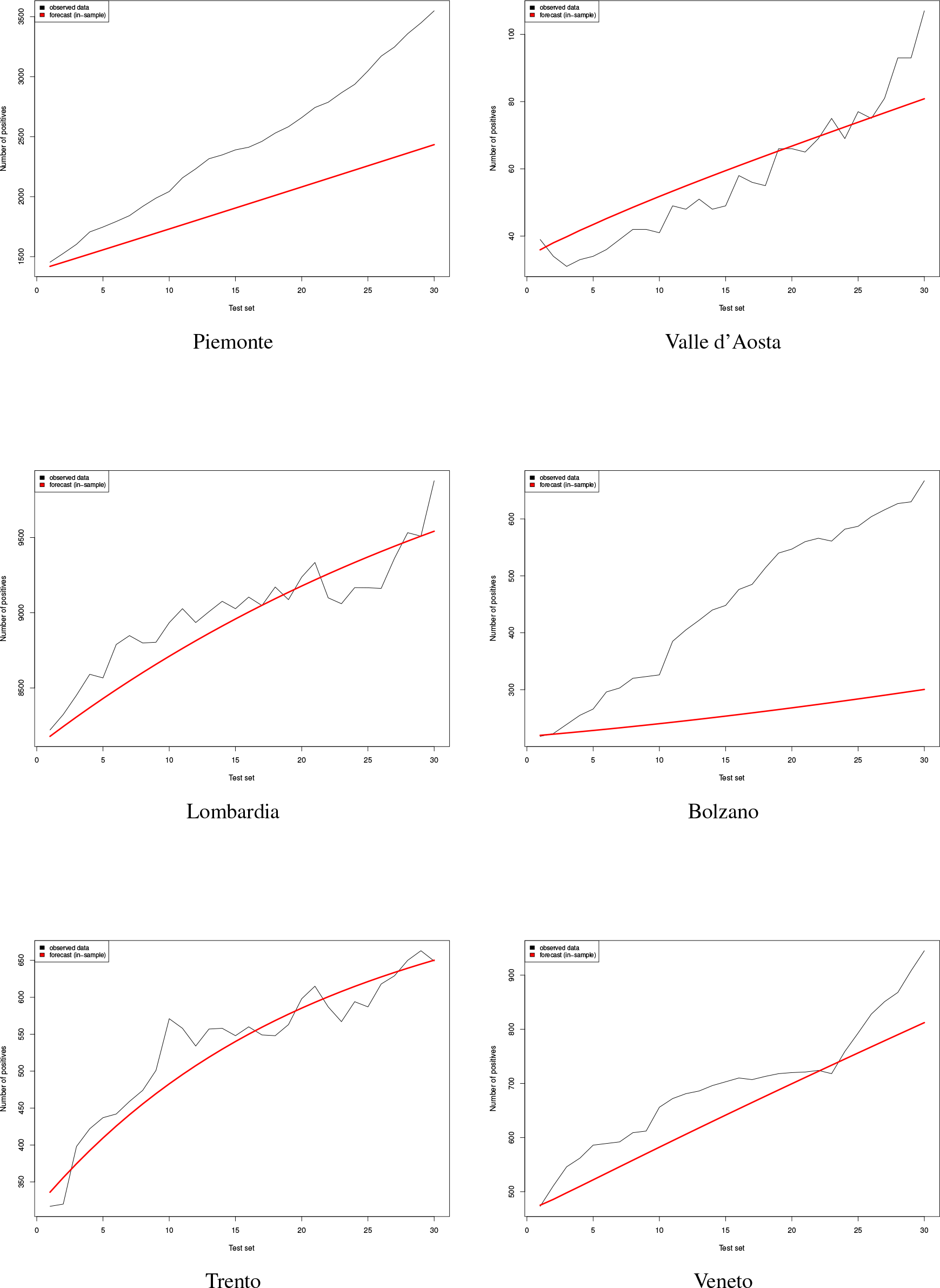
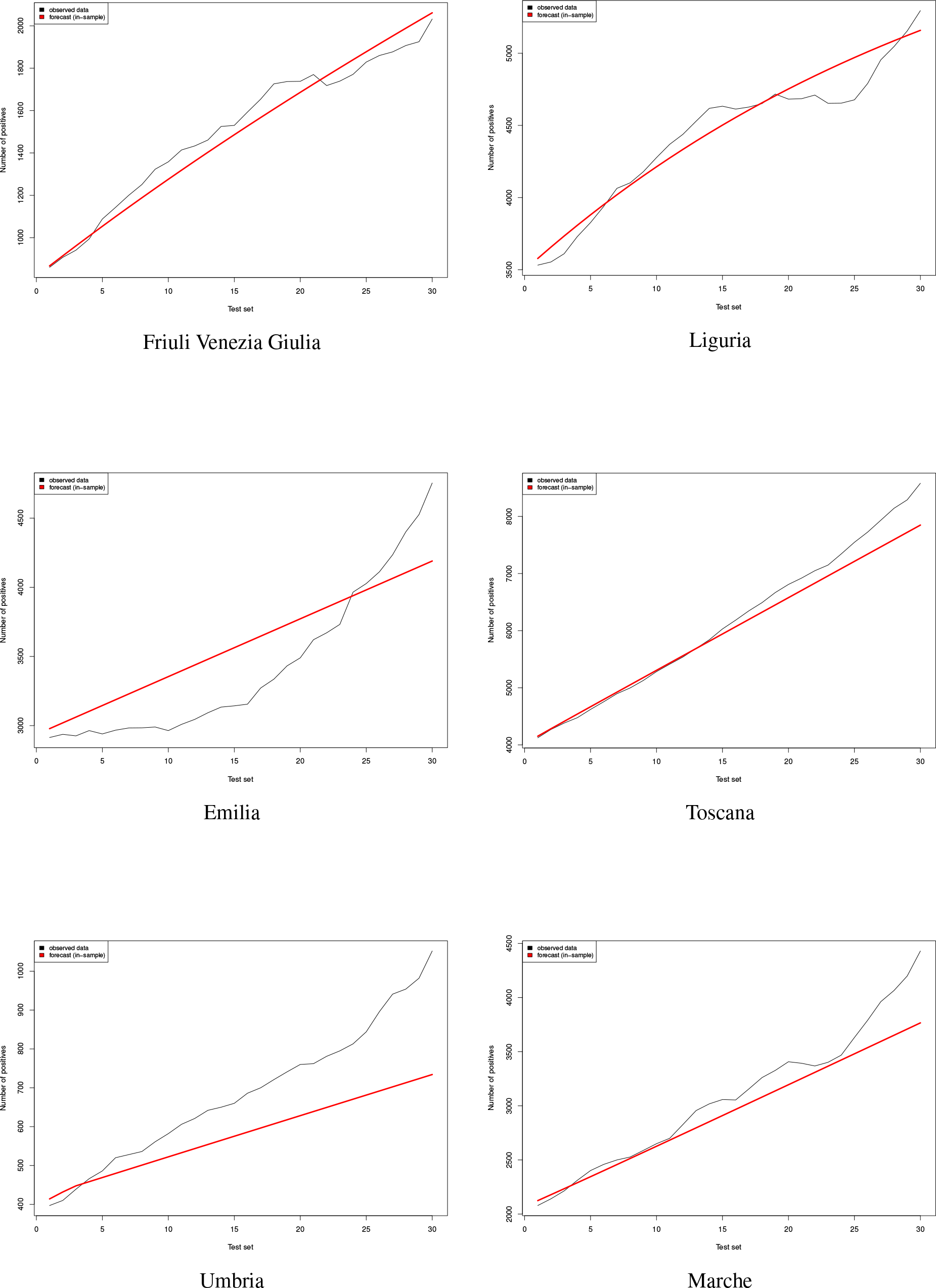
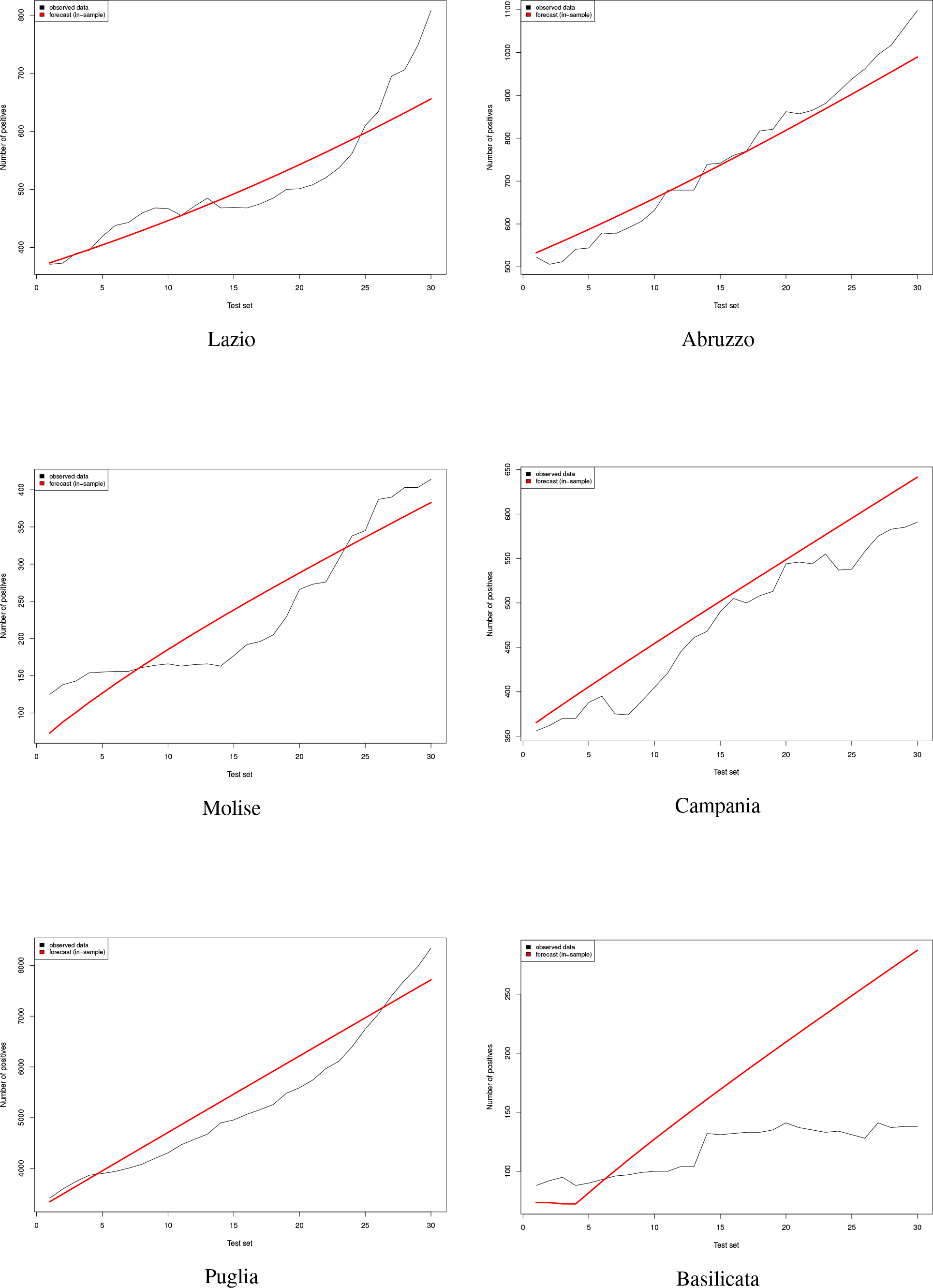
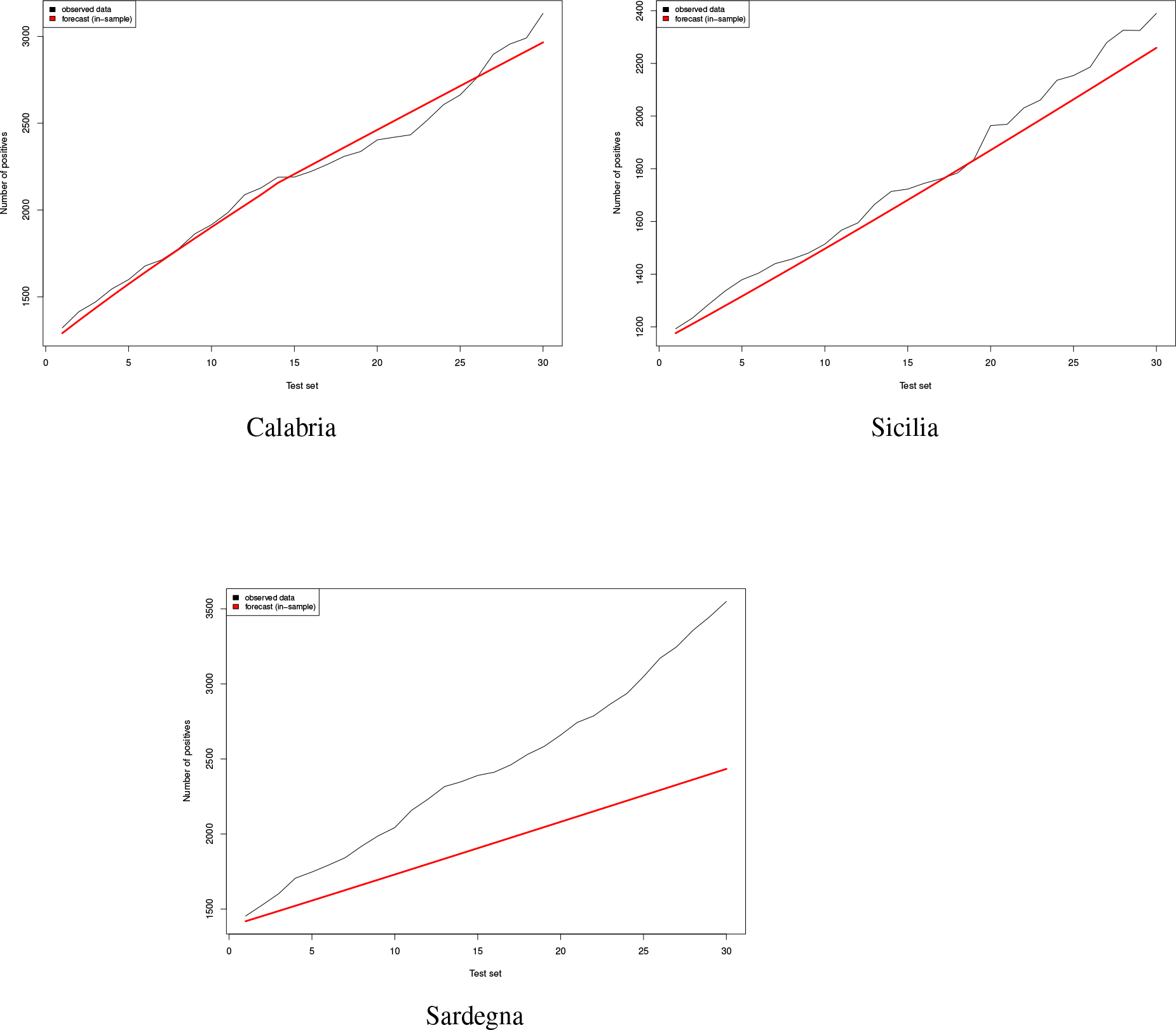
Goodness of fit of the out-of-sample predictions (the black and red lines refer respectively to the actual data (the set **𝒪**^test^) and the related predictions. Prediction window H = 30 days

## 11. Appendix B

**Fig. 6:**
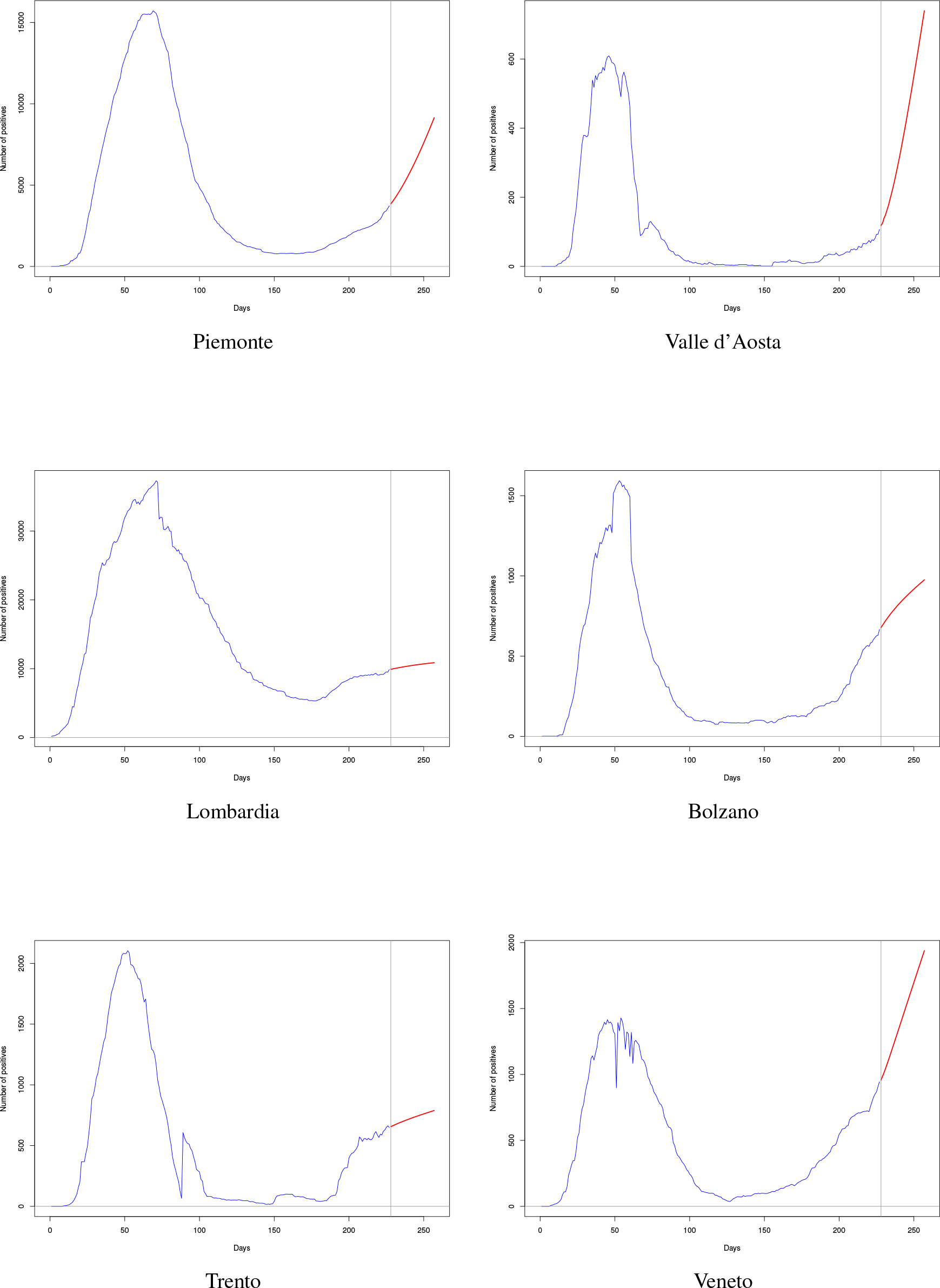
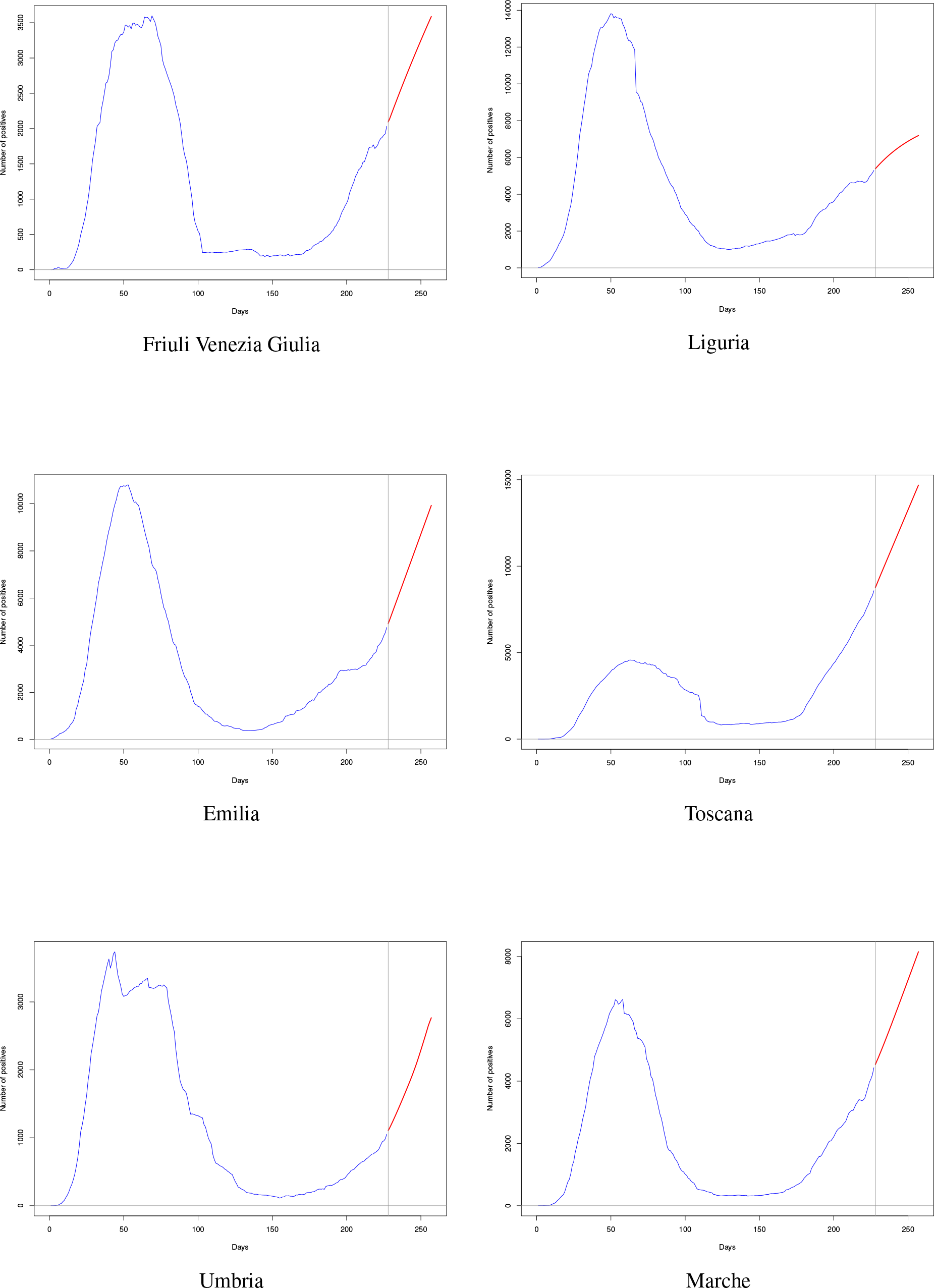
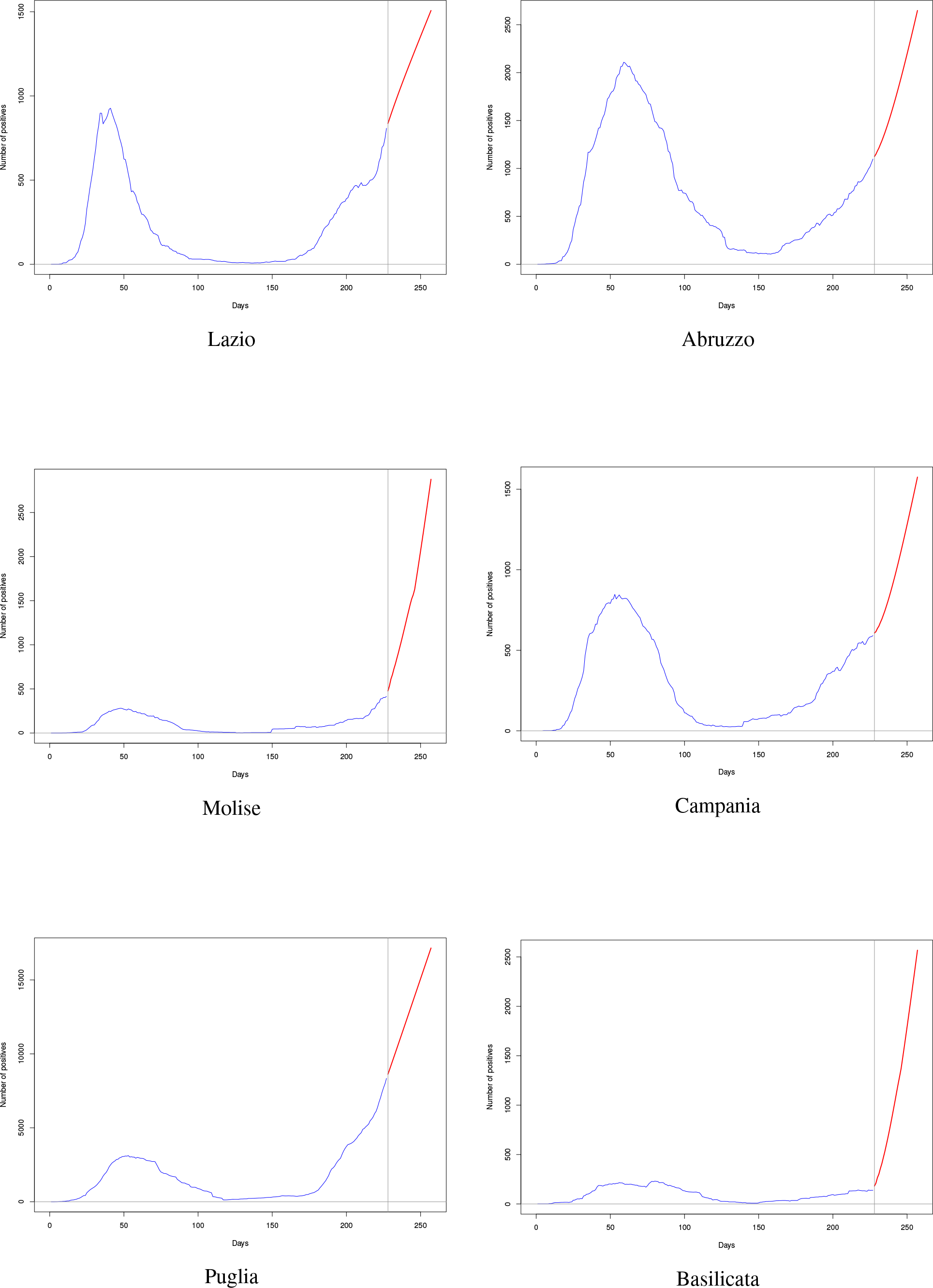
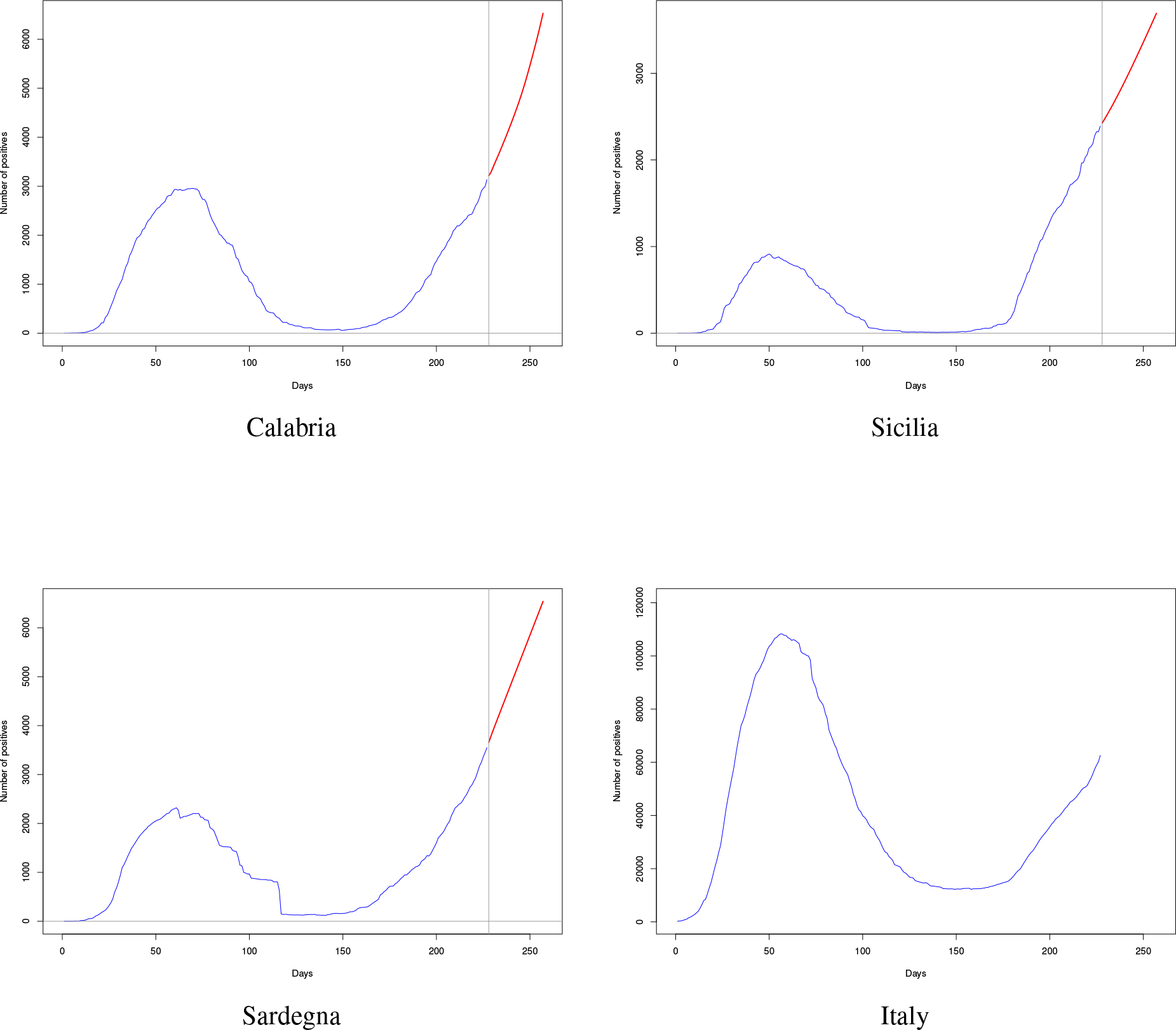
Out of sample pure forecasts. The Black lines refer to the actual data (the set **𝒪**) where the predictions for the horizon *H* = 3*O* days (the set **𝒪**^*fore*^) are in red

## Notes

### Competing Interest Statement

The authors have declared no competing interest.

### Funding Statement

No funding

